# Self-Reported Use of COVID-19 Immunologic Test Results to Inform Decisions About Daily Activities and COVID-19 Vaccination

**DOI:** 10.1101/2022.07.01.22277108

**Authors:** Miao Jiang, Nicholas K. Akers, Darcy B. Gill, Benjamin Eckhert, Emily Svejnoha, Harlan Robins

## Abstract

**Importance:** Despite widespread use of clinical diagnostic tests to assess prior exposure to SARS-CoV-2, limited evidence exists regarding how test results affect patient behaviors and decision-making.

**Objective:** To understand the rationale behind ordering diagnostic T-cell receptor (TCR) immunosequencing for assessment of prior SARS-CoV-2 infection and evaluate how test results affect patient behaviors, including day-to-day activities and decisions about vaccination.

**Design:** Mandatory demographic information and clinical characteristics were collected for all individuals ordering T-Detect™ COVID. Study participants completed a one-time survey that included additional questions about demographics and clinical characteristics, relevant interactions with healthcare providers, reasons for ordering diagnostic TCR immunosequencing, and the utility of test results.

**Setting:** US participants ordering T-Detect COVID between February 2021 and March 2022.

**Participants:** Of the 806 individuals who underwent diagnostic TCR immunosequencing, provided informed consent, and were sent the email survey, 718 completed the survey (response rate, 89.1%). At the time of receiving the test report, 25.5% of participants had been vaccinated against COVID-19, 29.7% reported a previous COVID-19 infection, and 25.6% were immunocompromised.

**Main Outcome(s) and Measure(s):** Patient demographics and clinical characteristics were reported using descriptive statistics. Additional analyses explored trends in reported data over time and evaluated reasons for ordering diagnostic TCR immunosequencing and behaviors among participant subgroups (vaccinated or unvaccinated individuals and those with positive or negative test results). Logistic regression analysis evaluated factors that increased the likelihood of post-test vaccination.

**Results:** Study participants ordered diagnostic TCR immunosequencing to understand their health status (55.0%) and to inform decision-making about daily activities (43.6%) and vaccination (38.3%). Most participants (92.1%) ordered diagnostic TCR immunosequencing for themselves without consulting their physician. Testing negative for prior SARS-CoV-2 infection was associated with increased likelihood of subsequent COVID-19 vaccination (31.0% vs 6.9%; median time to vaccination, 17.0 days vs 47.5 days), which was confirmed by logistic regression analysis.

**Conclusions and Relevance:** This report presents patient-reported clinical utility of a commercial COVID-19 assay based on an immune response readout. Our findings suggest that participants used diagnostic TCR immunosequencing results to inform decisions about daily activities and COVID-19 vaccination.

**Trial Registration:** Not applicable.

**KEY POINTS:**

- We aimed to understand the factors driving immunologic testing for SARS-CoV-2 and characterize the actions and decisions spurred by test results.
- Results of this study suggest that individuals frequently ordered immunologic testing for themselves to understand their health status and to inform decision-making about daily activities and vaccination.
- Among unvaccinated participants, testing negative for prior SARS-CoV-2 infection was associated with increased likelihood of undergoing vaccination and shorter time to vaccination.
- This study provides the first real-world evidence of patient-perceived utility of a COVID-19 immunologic test for decision-making related to vaccination and lifestyle.

## INTRODUCTION

Infection- or vaccination-mediated exposure to bacterial or viral antigens induces humoral (antibody-driven) and cellular (T-cell-driven) adaptive immune responses of varying strength and duration [1,2]. Diagnostic tests probing these immune responses, as well as assays that directly detect SARS-CoV-2, have played an important role in the public health response to COVID-19. At the community level, immunologic testing has been used to screen convalescent plasma for therapeutic use and to assess COVID-19 prevalence to inform strategies for pandemic management and prioritization of vaccine allocation [3–5]. At the individual level, immunologic testing for SARS-CoV-2 can allow patients to better understand their health status by identifying a previous infection [3]. For example, evidence of a previously undetected infection may be important for managing “long COVID” or multisystem inflammatory syndrome in children (MIS-C) [6,7].

Currently, serologic blood tests are the most common modality for detecting past SARS-CoV-2 infection [4,8]. However, utility of these antibody-based tests may be reduced in some individuals due to low or absent antibody titers [9–12]. Furthermore, serologic testing is not informative for some patients, such as those undergoing iatrogenic B-cell depletion or with primary humoral immunodeficiencies [13,14].

In addition to humoral responses, mounting evidence suggests that T-cell responses to SARS-CoV-2 contribute to early, durable protection from COVID-19 and may attenuate disease severity [10,15–19]. Greater conservation of T-cell epitopes across SARS-CoV-2 variants support a broader range of antigen recognition in emerging variants compared to antibody responses [20–22]. Capitalizing on these data, T-cell-based diagnostics have emerged as another strategy to probe the adaptive immune response for identifying prior SARS-CoV-2 infection [23–25]. In March 2021, T-Detect™ COVID became the first T-cell– based COVID-19 diagnostic to receive Emergency Use Authorization from the US Food and Drug Administration (FDA). This consumer-directed test identifies prior SARS-CoV-2 infection by using next-generation sequencing of whole blood samples to assess enrichment of SARS-CoV-2–specific T-cell receptors (TCRs) among the TCR repertoire [23,26–29].

Despite widespread use of immune-based COVID-19 diagnostics, our understanding of why patients choose to undergo testing and how test results affect their subsequent behaviors and clinical decision-making is incomplete. One small study of industrial workers suggested that implementation of serologic testing in an industrial environment with efficient protective measures may reduce fear without eliciting non-adherence to established safety protocols [30]. However, results of a recent randomized clinical trial suggest that patients using at-home COVID-19 self-testing were not likely to follow Centers for Disease Control and Prevention (CDC) recommendations for quarantine based on test results [31]. To gain insight into patient-perceived utility of COVID-19 diagnostics, we surveyed individuals who had ordered T-Detect COVID. We aimed to characterize the demographics of the patient population, better understand patients’ rationales for testing, and evaluate how test results affected subsequent behaviors, including day-to-day activities and decisions about vaccination. Results of this study provide real-world evidence demonstrating the value of immunologic testing in informing patient behaviors.

## METHODS

### Study ethics and approval

Survey data were collected pursuant to an Institutional Review Board-approved clinical study protocol (PRO-00854, WIRB# 20210171). Written informed consent was obtained from all participants.

### Assay ordering

In February 2021, the T-Detect COVID test became available for consumers to order. The online ordering process captured responses to a number of questions that addressed assay-specific eligibility criteria, as well as data required by Health and Human Services (HHS) mandatory reporting guidelines applicable to all SARS-CoV-2 immunologic tests [32]. Topics included demographic information (age, sex, race/ethnicity, geography) and medical history for select conditions associated with increased risk for severe COVID-19 (chronic conditions, obesity/overweight, immunocompromised status, tobacco use, heart conditions, neurologic conditions). The form also asked about prior COVID-19 testing history and whether the individual was experiencing specific symptoms associated with COVID-19. The order form included an option to opt-in to future research opportunities with Adaptive Biotechnologies. After physician review, individuals meeting criteria for eligibility received a prescription for the assay, and blood draws were conducted either via mobile phlebotomy or at a testing laboratory. Blood samples were sent to the Adaptive Biotechnologies central laboratory for processing. Results were shared through an online portal.

### Participant enrollment

Starting in October 2021, a survey was conducted among individuals who were tested with the T-Detect COVID assay. Participants who provided written informed consent answered a one-time questionnaire that included demographics; COVID-19 testing, vaccination, and infection history; and medical history (**Supplemental File**). Participants who reported a current autoimmune condition or cancer diagnosis and those who reported taking a medication that weakens the immune system were considered immunocompromised. The survey also assessed the clinical utility of diagnostic TCR immunosequencing with questions about who placed the order, rationale for ordering, how results were used, and perceived assay utility.

### Statistical analysis

Participant demographics, clinical characteristics, and self-reported utility of test results were summarized using descriptive statistics. Trends in test ordering, rationales for testing, and post-test behaviors over time were compared using graphical representations. Vaccination uptake was analyzed among previously unvaccinated individuals using descriptive statistics and presented graphically over time for the overall population of unvaccinated individuals and for subgroups defined by whether the test result was used to inform vaccine decision. Differences in vaccination rates between individuals with positive versus negative test results were assessed using Fisher’s Exact Test (FET). Median time to vaccination from test report delivery date was calculated. Logistic regression analysis was also performed to estimate the probability of post-test vaccination among individuals who were unvaccinated at the time of the report, adjusting for relevant demographic and clinical characteristics. Data analyses were conducted using R version 4.1.1. *P* values less than 0.05 were considered statistically significant.

## RESULTS

### Demographic and clinical characteristics of the study population

As of April 2022, among the approximately 30,000 individuals who underwent diagnostic TCR immunosequencing, 719 individuals completed the clinical utility survey (**Supplemental Figure 1**); 1 participant was excluded due to a data input error (response rate, 89.1% [718/806]). Timing of test orders among survey respondents was generally consistent with that of the testing population, with a peak in August and September of 2021 corresponding to the SARS-CoV-2 Delta wave in the US [33,34] (**Figure 1A**).

**Figure 1.**
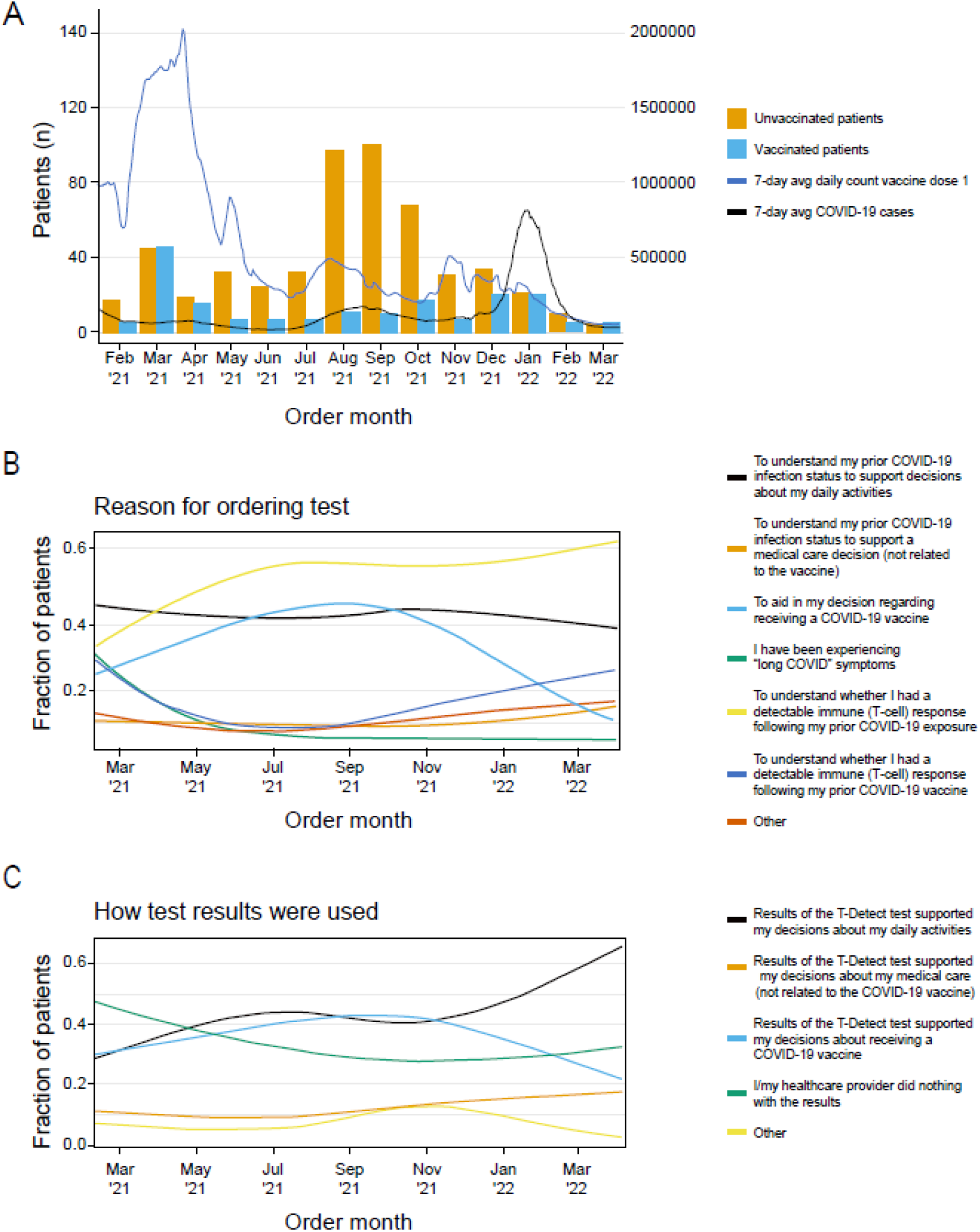
Trends in ordering diagnostic TCR immunosequencing and patient-reported utility of results over time. A. Proportion of vaccinated and unvaccinated participants ordering testing by month. Seven-day moving averages of COVID-19 cases and first COVID-19 vaccine doses were obtained from the CDC COVID Data Tracker (https://covid.cdc.gov/covid-data-tracker/#datatracker-home). B., C. Patient-reported reasons for ordering testing (B) and utility of test results (C) varied over time.

Survey respondents were predominantly white (89.7%) and non-Hispanic (89.7%); more than half were female (54.3%). Approximately one-tenth of participants were age 65 or older, and one-fourth were immunocompromised (**Table 1**). Demographics, geography, and reported comorbidities of the study population were consistent with those of patients who ordered the test, although survey respondents skewed slightly younger (>65 years: survey, 9.7%; all patients: 12.0%; **Supplemental Table 1**).

**Table 1.** Participant Demographics and Clinical Characteristics

|  | Survey participants<br>(n=718) |
| --- | --- |
| **Age, n (%)** |  |
| ≥65 years | 70 (9.7) |
| **Sex, n (%)** |  |
| Female | 390 (54.3) |
| Male | 328 (45.7) |
| **Ethnicity, n (%)** |  |
| Hispanic or Latino | 42 (5.8) |
| Non-Hispanic or Latino | 644 (89.7) |
| Unknown | 32 (4.5) |
| **Race, n (%)** |  |
| American Indian or Alaska native | 7 (1.0) |
| Asian | 33 (4.5) |
| Black or African American | 6 (0.8) |
| White | 644 (89.7) |
| Unknown | 28 (3.9) |
| **Geography, n (%)** |  |
| Midwest | 127 (17.7) |
| Northeast | 142 (19.8) |
| South | 183 (25.5) |
| West | 266 (37.0) |
| **First COVID-19 test of any kind, n (%)** | 122 (17.0) |
| **T-Detect COVID result, n (%)** |  |
| Negative | 363 (50.6) |
| Positive | 355 (49.4) |
| **Immunocompromised condition,<sup>a</sup> n (%)** | 184 (25.6) |
| **COVID-19 status at time of report, n (%)** |  |
| No known prior infection | 505 (70.3) |
| Infection date known | 193 (26.9) |
| Infection date unknown | 20 (2.8) |
| **COVID-19 vaccination status at time of report, n (%)** |  |
| Unvaccinated | 535 (74.5) |
| Vaccinated | 183 (25.5) |

TCR, T-cell receptor.
<sup>a</sup>Immunocompromised individuals were defined as those with autoimmune conditions, cancer, or taking medications that weaken the immune system.

At the time of receiving their test report, 25.5% of participants reported that they had been vaccinated against COVID-19, and 29.7% reported a previous COVID-19 diagnosis. Survey respondents were less likely to have had prior COVID-19 testing (17.0% vs 23.7% overall) and had a higher rate of T-Detect COVID positivity (indicating prior SARS-CoV-2 infection) compared to the broader population of patients who underwent testing (49.4% vs 28.1% overall, **Supplemental Table 1**).

### Patients used diagnostic TCR immunosequencing results to inform decision-making related to COVID-19

Respondents reported that the most common reasons for ordering diagnostic TCR immunosequencing were to understand whether the individual had a detectable T-cell immune response following prior COVID-19 exposure (55.0%; 395/718), to support decisions about daily activities (43.6%; 313/718), and to aid in decision-making about receiving a COVID-19 vaccine (38.3%; 275/718) (**Table 2**). In addition, among the smaller group of vaccinated individuals, understanding the immune response after COVID-19 vaccination was a commonly reported rationale (44.9%; 70/156). Other reasons for testing included understanding prior COVID-19 infection status to make a medical care decision not related to vaccination (10.2%; 73/718) and gaining insight into whether current “long COVID” symptoms may be due to an undiagnosed prior infection (8.8%; 63/718). With regard to utility, 41.4% (297/718) of respondents reported that the test result supported their decisions about daily activities, and 38.3% (275/718) reported that their results supported their decision about receiving a COVID-19 vaccine.

**Table 2.** Patient-Reported Clinical Utility of Diagnostic TCR Immunosequencing

|  | All survey participants<br>(n=718) |
| --- | --- |
| **Role of the healthcare provider in ordering test, n (%)** |  |
| Healthcare provider recommended | 23 (3.2) |
| Patient decided alone | 661 (92.1) |
| Patient and provider decided together | 34 (4.7) |
| **Rationale for ordering test, n (%)** |  |
| To understand my prior COVID-19 infection status to support decisions about my daily activities | 313 (43.6) |
| To understand my prior COVID-19 infection status to support a medical care decision (not related to the vaccine) | 73 (10.2) |
| To aid in my decision regarding receiving a COVID-19 vaccine | 275 (38.3) |
| I have been experiencing “long COVID” symptoms and have not previously been positively diagnosed with COVID-19 or to determine whether my long COVID symptoms are related to a past infection that has not previously been positively diagnosed | 63 (8.8) |
| To understand whether I had a detectable immune (T-cell) response following my prior COVID-19 exposure | 395 (55.0) |
| To understand whether I had a detectable immune (T-cell) response following my prior COVID-19 vaccine | 104 (14.5) |
| Other | 76 (10.6) |
| **Discussed test results with healthcare provider, n (%)** |  |
| Yes | 158 (22.0) |
| Have not, but plan to do so | 154 (21.4) |
| Have not, and do not plan to do so | 406 (56.5) |
| **Utility of test results, n (%)** |  |
| Results of the T-Detect test supported my decisions about my daily activities | 297 (41.4) |
| Results of the T-Detect test supported my decisions about my medical care (not related to the COVID-19 vaccine) | 84 (11.7) |
| Results of the T-Detect test supported my decisions about receiving a COVID-19 vaccine | 275 (38.3) |
| I/my healthcare provider did nothing with the results | 233 (32.5) |
| Other | 61 (8.5) |
| **Rating of perceived utility, n (%)** |  |
| 5: Extremely useful | 271 (37.7) |
| 4: Very useful | 182 (25.3) |
| 3: Somewhat useful | 160 (22.3) |
| 2: Slightly useful | 53 (7.4) |
| 1: Not at all useful | 52 (7.2) |

To evaluate time-dependent trends in rationale and test utility, we plotted the data longitudinally. Reasons for undergoing testing remained mostly constant over time (**Figure 1B**), although the number of participants interested in understanding their immune response after exposure increased, consistent with the growing number of infections in the US over the same period [34]. The most dynamic reason for ordering the test was to aid in decision-making regarding receiving a COVID-19 vaccine, which peaked in August to September 2021 and dropped substantially in early 2022 (**Figure 1B**). In alignment with participants’ rationale for undergoing testing to inform decisions about vaccination, the number of patients who reported using test results to support their decision-making about COVID-19 vaccination peaked in October 2021 (**Figure 1C**).

Many patients reported using their test results to aid in decision-making related to daily activities (41.4%; 297/718) or vaccination (38.3%; 275/718), but they may have been making these decisions without guidance from their healthcare providers. Most participants (92.1%; 661/718) ordered diagnostic TCR immunosequencing for themselves without consulting their physician. After receiving their results, only 22.0% (158/718) discussed their results with their healthcare provider, although another 21.4% (154/718) planned to do so in the future (**Table 2**).

### Patients who received negative test results were more likely to undergo COVID-19 vaccination

To better understand how patients used immunologic test results to inform decisions about vaccination, we examined rates of vaccination among those who were not vaccinated prior to receiving their report. Among these individuals, the overall rate of vaccination was higher for patients who received a negative result compared to those who tested positive for prior SARS-CoV-2 infection (31.0% [94/303] vs 6.9% [16/232]; **Figure 2A**). The interval between receiving test results and subsequent vaccination was also examined, revealing that participants who received a negative result were also vaccinated sooner after receiving their results (median time to vaccination, 17.0 days vs 47.5 days; **Figure 2B**).

**Figure 2.**
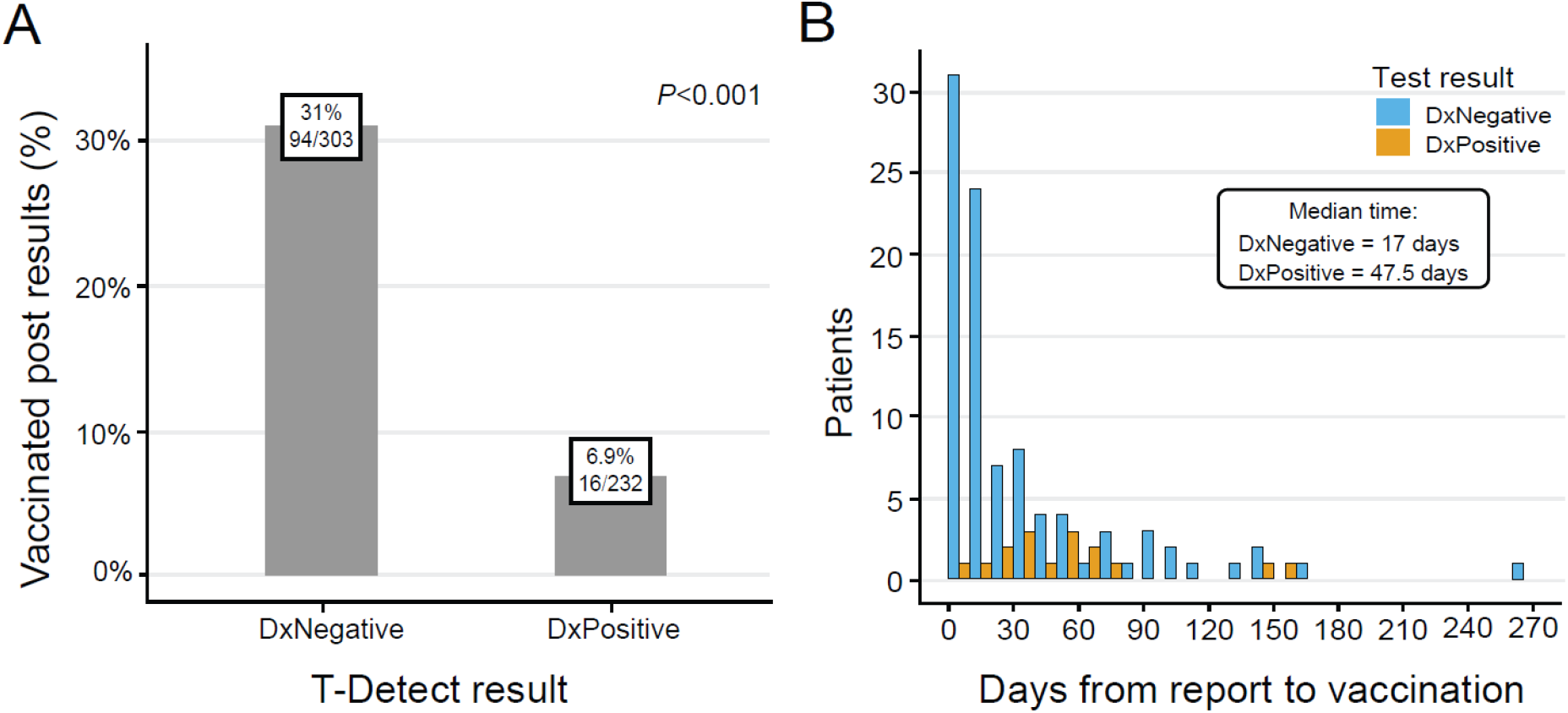
Vaccination status and timing stratified by diagnostic TCR immunosequencing COVID test results. A. Percentage of patients vaccinated after receiving test results stratified by positive or negative result overlayed with 7-day moving average of COVID-19 cases and people receiving at least 1 dose of vaccine per the CDC COVID Data Tracker site (https://covid.cdc.gov/covid-data-tracker/#datatracker-home). B. Time to vaccination after receiving test results stratified by positive or negative result. Dx, diagnosis; FET, Fisher’s exact test.

To further evaluate the relationship between the decision to undergo vaccination and the test result, we examined trends in vaccination over time, stratified by test result and whether participants reported using test results to decide about vaccination. The rate of vaccination decreased over time across all subgroups. However, the group of participants who tested negative and reported using their results to inform vaccination decisions had the highest rate of vaccination, while those who tested positive and reported using their results to inform vaccination decisions had the lowest rate of vaccination. In contrast, participants who did not report using their test results to inform vaccination exhibited no apparent difference in vaccination rates over time, regardless of whether they tested negative or positive. This evidence supports the conclusion that testing negative for prior SARS-CoV-2 infection was associated with a higher likelihood of undergoing COVID-19 vaccination among individuals who sought out testing to inform their decision. However, it is also important to note that timing of the test was a key driving factor. Differences in vaccination rates between patients testing positive and negative were greatest in the earliest months after the test became available (March 2021–October 2021) and diminished over time, with low rates of vaccination observed among both groups in 2022 (**Figure 3**).

**Figure 3.**
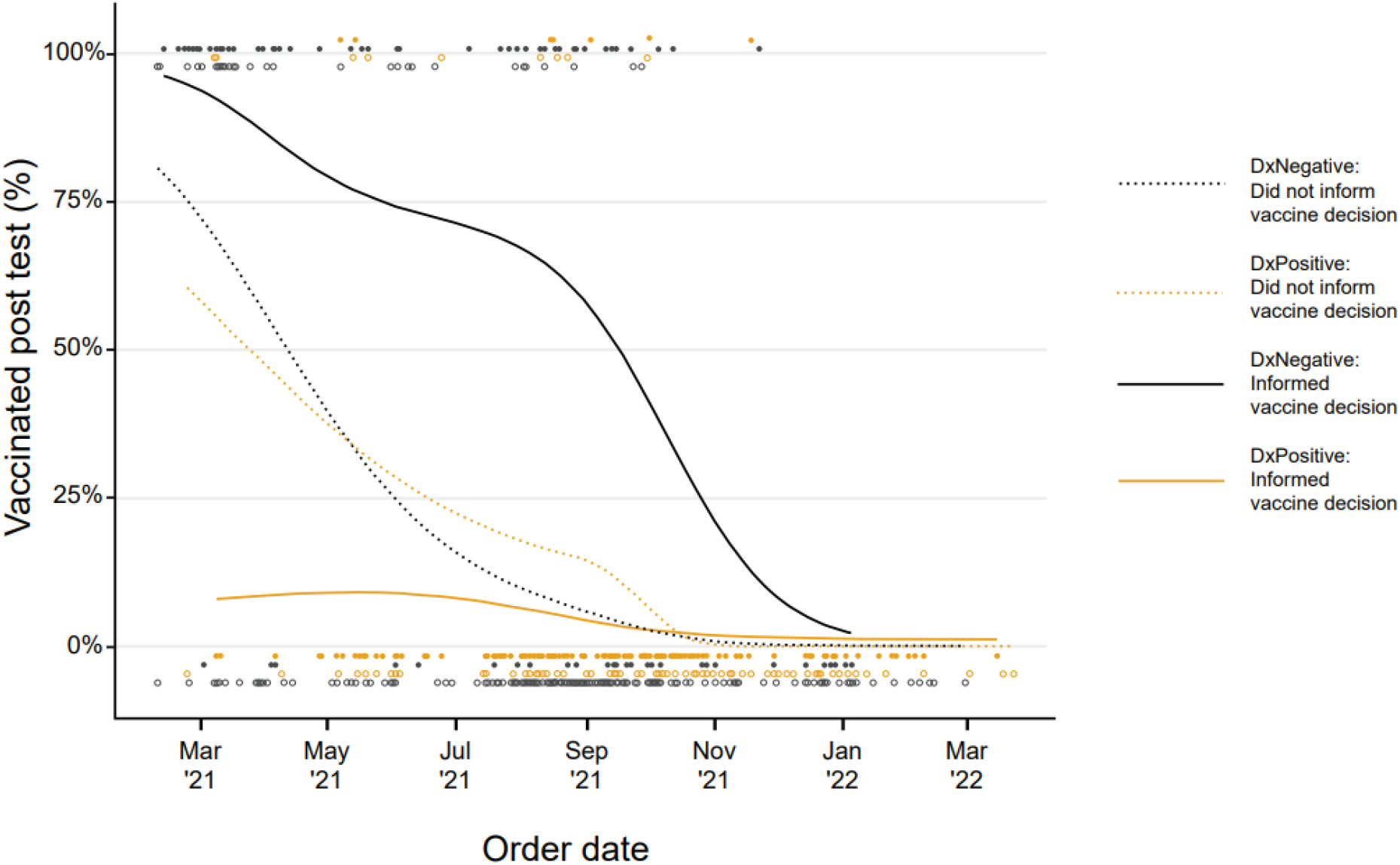
Use of diagnostic TCR immunosequencing results to inform the decisions about vaccination against COVID-19. The fraction of patients who self-reported using test results to inform the decision to undergo COVID-19 vaccination was stratified by the test result. The magnitude of the difference varied over time, with the largest differences observed in March 2021 to October 2021. Dx, diagnosis.

To statistically evaluate factors that contributed to the decision to undergo vaccination, we performed logistic regression analysis. The results were consistent with graphical trends. The only factors that significantly increased the likelihood of vaccination were negative test results (odds ratio [OR], 13.8; 95% confidence interval [CI], 5.4–35.7), receipt of report in the first 6 months of the study period (OR, 9.8; 95% CI, 5.5–17.6), and self-reported use of testing to inform vaccination decision (OR, 4.3; 95% CI, 2.3–7.8) (**Supplemental Table 2**). Collectively, these results suggest that participants may have used negative results from diagnostic TCR immunosequencing to inform the decision to get vaccinated against COVID-19.

## DISCUSSION

This study analyzed patient-reported outcomes among those who ordered a novel, T-cell-based, commercial COVID-19 immune-response diagnostic test. The results showed that the most common reasons for ordering the test were to understand T-cell immune response status after exposure to COVID-19, to inform decisions about daily activities, and to assist in decision-making about receiving a COVID-19 vaccine. Given that most participants ordered the test for themselves without consulting their physician, many were likely making decisions related to daily activities and vaccination without guidance from their healthcare providers. Consistent with these results, removal of many governmental mitigation strategies left individuals to rely on their own risk assessments to guide their daily activities during the pandemic [35], and the emphasis on large-scale vaccination centers has decreased physician involvement in vaccination decisions [36,37].

The utility of immunologic testing to inform individual risk is limited [4], as ongoing emergence of new variants has hampered efforts to define a clear correlate of protection for COVID-19 [38,39]. Therefore, FDA does not recommend that immunologic testing be used as a measure of immunity or protection from COVID-19 infection, and immunologic tests that have been granted Emergency Use Authorization have not been evaluated to assess protection provided by the immune response to vaccination or infection [11]. Similarly, the World Health Organization (WHO) has recommended against the use of immunologic tests for so called “immunity passports” to enable international travel [40]. However, prior to this study, there were no published reports, backed by real-world evidence, on whether people taking these immunologic tests are adhering to FDA and WHO recommendations against using test results to infer immunity, thereby informing vaccination and lifestyle decisions [11,41,42].

Our findings also indicate that patients are potentially using negative results to inform the decision to get vaccinated. Unvaccinated participants who tested negative for prior COVID-19 were more likely to report subsequent vaccination (31.0% vs 6.9%) and sooner vaccination (median time to vaccination, 17 days vs 47.5 days) compared to those who tested positive. These results are consistent with surveys indicating that previous COVID infection influences attitudes towards vaccination [43–45]. However, as vaccination is a complex multifaceted decision involving both personal and societal factors, these findings have to be interpreted in the broader context. This complexity is reflected in the logistic regression analysis, where in addition to testing negative, receipt of report in the first 6 months of the study period and self-reported use of testing to inform vaccination decisions also significantly increased the likelihood of post-test vaccination. Belief in vaccine efficacy is strongly correlated with vaccine acceptance [46,47], so reports of decreased protection against infection and transmission with viral variants, followed by evidence of reduced disease severity with the Omicron variant [48,49] may have contributed to vaccine hesitancy and decreased rates of vaccination among these individuals over time.

This study has several limitations. First, generalizability of the study results may be restricted as it involved a small number of participants who were not fully representative of the general population in a few aspects. For example, survey participants tended to be younger and predominantly white and were less likely to report being obese, having chronic medical conditions, or smoking compared to the general population [50–52] (**Supplementary Table 1**). Although income was not asked in the survey, given the self-pay nature of the test, we assume that the study population is likely weighted toward individuals from higher socioeconomic strata compared to the overall US population, similar to surveys of users of at-home COVID-19 tests [53]. Thus, the changes in behavior reported by participants in this study may not apply to individuals from other demographic strata. Second, no direct causality of the test result for subsequent vaccine behavior can be demonstrated, as participants were not asked whether they were vaccinated because of their test result. However, examination of the interval between receiving test results and vaccination and of the trend in vaccination rate among those who indicated using test results to inform vaccine decision provides evidence supporting an association between test results and vaccination. Finally, the timing of this study reflects a unique period during which the interplay between vaccine availability, the emergence of different viral variants, and changes in public perception are complex. These factors may allow the study to only capture a snapshot in time that may not be replicated in the future.

Approval of multiple vaccines [54–57] and widespread availability through mass vaccination campaigns [58] has significantly reduced the global health burden of COVID-19 [59–61]. However, vaccine hesitancy has slowed down the scale of vaccination [62], and vaccination rates have decreased with approval of each additional booster [63]. Even before the COVID-19 pandemic, vaccination hesitancy was listed among the top 10 global health threats by the WHO [64], and evidence-based strategies, such as vaccinating hesitant patients through familiar providers [65], are needed to overcome this barrier across populations. However, the findings of this study suggest that a subset of individuals may be more likely to make decisions about vaccination without seeking guidance from their providers. Among this population, a negative COVID-19 immune response test result may represent a potential factor to increase vaccination uptake. Thus, in addition to informing individual decision-making, diagnostics probing humoral and cellular immunity are important for obtaining a more complete profile of immune status, and can support population-level public health policies related to timing and promotion of COVID-19 vaccine boosters, as well as future development of interventions based on antibody and T-cell responses.

## Supporting information

T-Detect COVID-19 Study Questionnaire

## Data Availability

Data requests may be submitted for consideration to Adaptive Biotechnologies Medical Information.

https://www.adaptivebiotech.com/medical-information-request/

## Author Contributions

Miao Jiang, PhD, and Nicholas A. Akers, PhD, had full access to all the data in the study and takes responsibility for the integrity of the data and the accuracy of the data analysis.

*Concept and design:* All authors.

*Acquisition, analysis, or interpretation of data:* All authors.

*Drafting of the manuscript:* All authors.

*Critical revision of the manuscript for important intellectual content:* All authors.

*Statistical analysis:* Miao Jiang and Nicholas A. Akers.

*Administrative, technical, or material support:* All authors.

*Supervision:* Jiang, Robins.

## Data sharing

Data requests may be submitted for consideration to Adaptive Biotechnologies Medical Information (https://www.adaptivebiotech.com/medical-information-request/).

## DATA SUPPLEMENT

**Supplemental Table 1.**
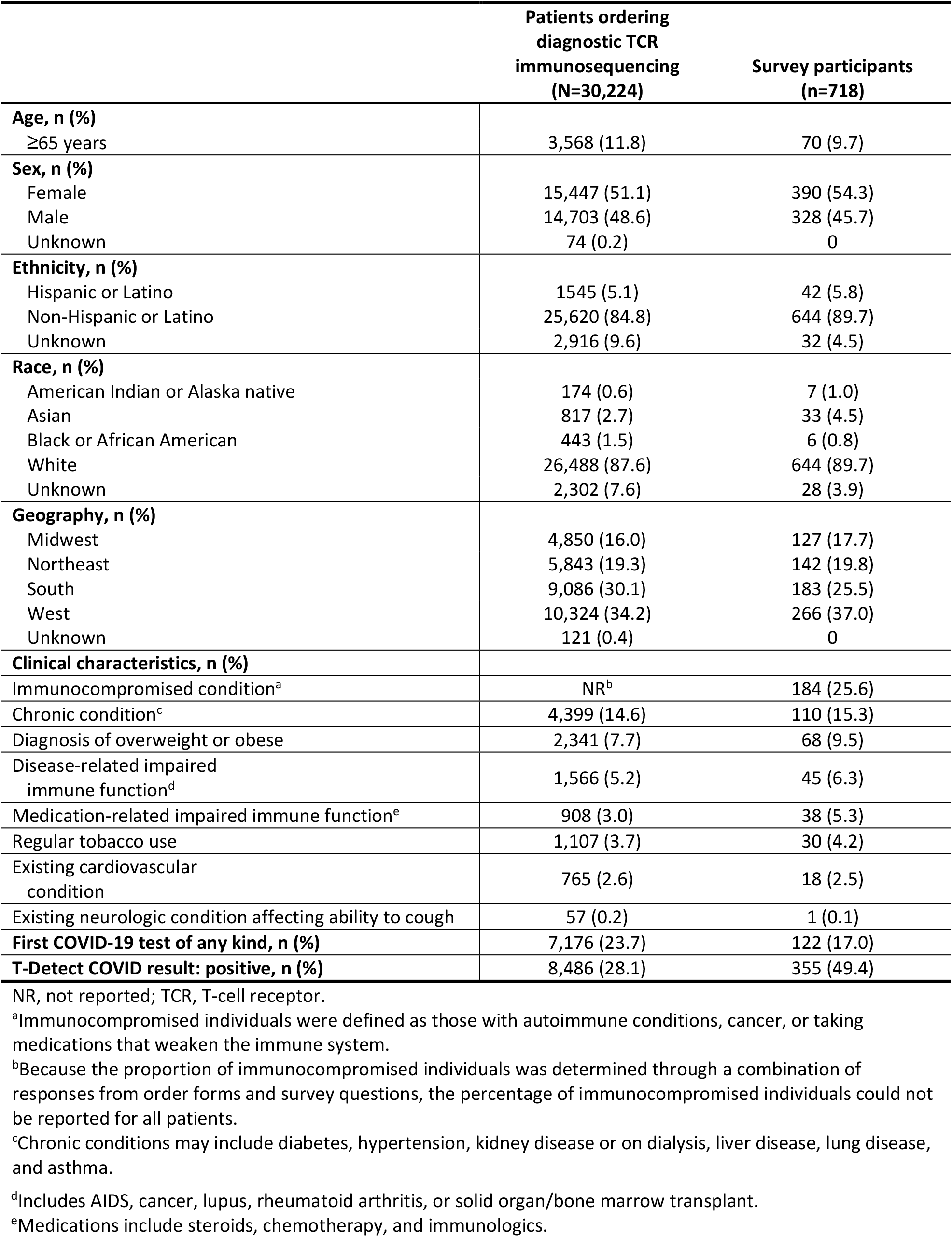
Comparison of demographics and clinical characteristics for survey participants and overall population ordering TCR immunosequencing.

|  | Patients ordering<br>diagnostic TCR<br>immunosequencing<br>(N=30,224) | Survey participants<br>(n=718) |
| --- | --- | --- |
| **Age, n (%)** |  |  |
| ≥65 years | 3,568 (11.8) | 70 (9.7) |
| **Sex, n (%)** |  |  |
| Female | 15,447 (51.1) | 390 (54.3) |
| Male | 14,703 (48.6) | 328 (45.7) |
| Unknown | 74 (0.2) | 0 |
| **Ethnicity, n (%)** |  |  |
| Hispanic or Latino | 1545 (5.1) | 42 (5.8) |
| Non-Hispanic or Latino | 25,620 (84.8) | 644 (89.7) |
| Unknown | 2,916 (9.6) | 32 (4.5) |
| **Race, n (%)** |  |  |
| American Indian or Alaska native | 174 (0.6) | 7 (1.0) |
| Asian | 817 (2.7) | 33 (4.5) |
| Black or African American | 443 (1.5) | 6 (0.8) |
| White | 26,488 (87.6) | 644 (89.7) |
| Unknown | 2,302 (7.6) | 28 (3.9) |
| **Geography, n (%)** |  |  |
| Midwest | 4,850 (16.0) | 127 (17.7) |
| Northeast | 5,843 (19.3) | 142 (19.8) |
| South | 9,086 (30.1) | 183 (25.5) |
| West | 10,324 (34.2) | 266 (37.0) |
| Unknown | 121 (0.4) | 0 |
| **Clinical characteristics, n (%)** |  |  |
| Immunocompromised condition<sup>a</sup> | NR<sup>b</sup> | 184 (25.6) |
| Chronic condition<sup>c</sup> | 4,399 (14.6) | 110 (15.3) |
| Diagnosis of overweight or obese | 2,341 (7.7) | 68 (9.5) |
| Disease-related impaired<br>immune function<sup>d</sup> | 1,566 (5.2) | 45 (6.3) |
| Medication-related impaired immune function<sup>e</sup> | 908 (3.0) | 38 (5.3) |
| Regular tobacco use | 1,107 (3.7) | 30 (4.2) |
| Existing cardiovascular<br>condition | 765 (2.6) | 18 (2.5) |
| Existing neurologic condition affecting ability to cough | 57 (0.2) | 1 (0.1) |
| **First COVID-19 test of any kind, n (%)** | 7,176 (23.7) | 122 (17.0) |
| **T-Detect COVID result: positive, n (%)** | 8,486 (28.1) | 355 (49.4) |

NR, not reported; TCR, T-cell receptor.
<sup>a</sup>Immunocompromised individuals were defined as those with autoimmune conditions, cancer, or taking medications that weaken the immune system.
<sup>b</sup>Because the proportion of immunocompromised individuals was determined through a combination of responses from order forms and survey questions, the percentage of immunocompromised individuals could not be reported for all patients.
<sup>c</sup>Chronic conditions may include diabetes, hypertension, kidney disease or on dialysis, liver disease, lung disease, and asthma.
<sup>d</sup>Includes AIDS, cancer, lupus, rheumatoid arthritis, or solid organ/bone marrow transplant.
<sup>e</sup>Medications include steroids, chemotherapy, and immunologics.

**Supplemental Table 2.** Logistic regression analysis of post-test vaccination among unvaccinated individuals (n=520), adjusted for demographic and clinical characteristics.

| Characteristic | Estimate | SE | Z value | P value | OR (95% CI) |
| --- | --- | --- | --- | --- | --- |
| **(Intercept)** | -4.615 | 0.626 | -7.376 | 0.000 |  |
| **Demographics** |  |  |  |  |  |
| Age: ≥65 years | 0.329 | 0.485 | 0.678 | 0.498 | 1.39 (0.54–3.59) |
| Sex: female | -0.387 | 0.277 | -1.394 | 0.163 | 0.68 (0.39–1.17) |
| Ethnicity: Hispanic or Latino | -1.040 | 0.662 | -1.571 | 0.116 | 0.35 (0.10–1.29) |
| **Geography** |  |  |  |  |  |
| Region: Midwest | -0.836 | 0.398 | -2.100 | 0.036 | 0.43 (0.20–0.95) |
| Region: Northeast | -0.468 | 0.370 | -1.265 | 0.206 | 0.63 (0.30–1.29) |
| Region: South | -0.818 | 0.352 | -2.328 | 0.020 | 0.44 (0.22–0.88) |
| **Clinical history** |  |  |  |  |  |
| Immunocompromised | -0.036 | 0.343 | -0.106 | 0.915 | 0.96 (0.49–1.89) |
| Prior COVID-19 diagnosis | 0.646 | 0.457 | 1.413 | 0.158 | 1.91 (0.78–4.67) |
| **Testing** |  |  |  |  |  |
| Test result: negative | 2.628 | 0.484 | 5.430 | <0.001 | **13.84 (5.36–35.73)** |
| Report received prior to August 31, 2021 | 2.286 | 0.298 | 7.675 | <0.001 | **9.83 (5.49–17.63)** |
| Self-reported use of testing to inform vaccination | 1.449 | 0.306 | 4.733 | <0.001 | **4.26 (2.34–7.76)** |

CI, confidence interval; OR, odds ratio; SE, standard error.

**Supplemental Figure 1.**
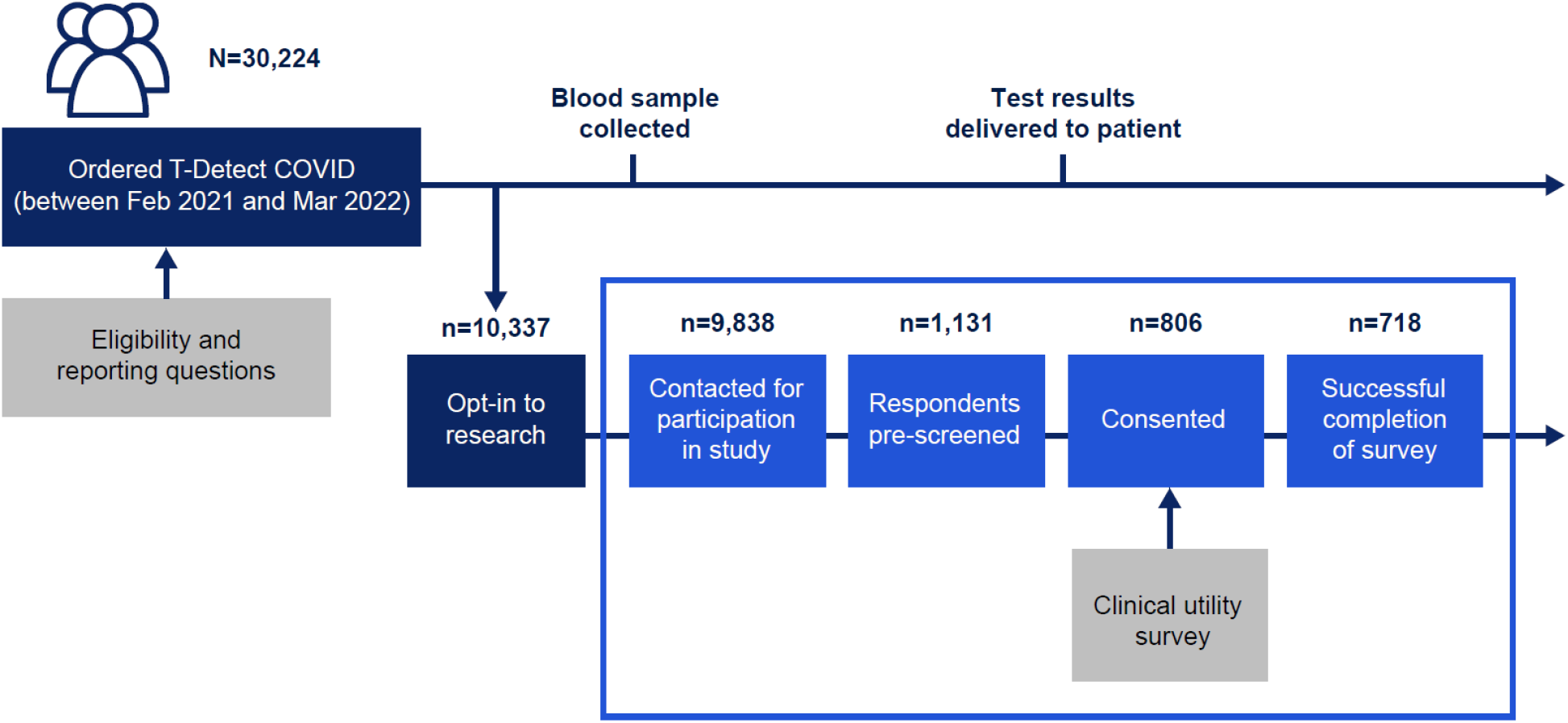
Study overview and patient allocation.

**Supplemental File. T-Detect COVID survey**.

