## Supplementary material for "Self-Reported Use of COVID-19 Immunologic Test Results to Inform Decisions About Daily Activities and COVID-19 Vaccination": T-Detect COVID-19 Study Questionnaire

Demographic Information

Q1. What is your biological sex?
□ Male
□ Female
□ Other

If Q1 = Other
Q2. Please describe your biological sex:
____________________________

Q3. What is your race/ethnicity? Select all that apply:
□ American Indian or Alaskan Native
□ Asian
□ Black or African American
□ Hispanic or Latino/a
□ Native Hawaiian or Other Pacific Islander
□ White
□ Other
□ Unknown

If Q3 = Other
Q4. Please describe your race/ethnicity:
____________________________

IRB Approved at the
Protocol Level
Sep 27, 2021

COVID-19 Vaccine Information

Q1. Have you received a vaccine against COVID-19?
□ Yes
□ No
If Q1 = No → T-Detect COVID-19 Test Information

If Q1 = Yes
Q2. How many times had you received a dose of a COVID-19 vaccine?
□ 1
□ 2
□ 3

Q2 will determine how many q's below the participant receives.
Q3. When did you receive your first dose of the vaccine (Month/Day/Year)?

______________
(MM/DD/YYYY)

Q4. When did you receive your second dose of the vaccine (Month/Day/Year)?

______________
(MM/DD/YYYY)

Q5. When did you receive your third dose (or booster) of the vaccine (Month/Day/Year)?

______________
(MM/DD/YYYY)

Q6. Which vaccine did you receive? The company name that developed the vaccine will be listed on the vaccine card you were given after you received it.
□ Pfizer-BioNTech
□ Moderna
□ Janssen (Johnson & Johnson)
□ AstraZeneca
□ Novavax
□ Other
□ Not sure

If Q6 = Other
Q7. Please list the other vaccine you received.

_________________________

IRB Approved at the
Protocol Level
Sep 27, 2021

T-Detect COVID-19 Test Information

Q1. When did you give your blood sample for the T-Detect COVID Test?

______________
(MM/DD/YYYY)

Q2. Were you experiencing symptoms of COVID-19 when you gave your blood sample for the T-Detect COVID Test?
□ Yes
□ No

If Q2=Yes
Q3. In the few days before you gave your blood sample for the T-Detect COVID test, had your symptoms gotten better, gotten worse or stayed the same?
□ Gotten better
□ Gotten worse
□ Stayed the same

Q4. Did you or your healthcare provider decide to order the T-Detect COVID Test?
□ My healthcare provider recommended the test
□ I wanted to order the test
□ I wanted to order the test and discussed it with my healthcare provider, and we decided together

If Q4 = I wanted to order the test or I wanted to order the test and discussed it with my healthcare provider, and we decided together
Q5. Why did you want to order the T-Detect COVID Test? (Select all that apply)
□ I wanted to understand my prior COVID infection status to support decisions about my daily activities.
□ I wanted to understand my prior COVID infection status to support a medical care decision (not related to the vaccine).
□ To aid in my decision regarding receiving a COVID vaccine
□ I have been experiencing "Long-COVID" symptoms and have not previously been positively diagnosed with COVID.
□ I wanted to understand whether I had a detectable immune (T-cell) response following my prior COVID exposure.
□ I wanted to understand whether I had a detectable immune (T-cell) response following my prior COVID vaccine.
□ Other

If Q5 = Other
Q6. Please describe the other reason you decided to order the T-Detect COVID Test?

_________________________

Q7. Did you discuss the results of the T-Detect COVID test with your healthcare provider?
□ Yes

□ I have not discussed the results with my healthcare provider yet, but I do plan to.
□ I have not discussed the results with my healthcare provider and do not plan to.

Q8. What did you do with the results of the T-Detect COVID test? (Select all that apply)
□ Results of the T-Detect test supported my decisions about my daily activities.
□ Results of the T-Detect test supported my decisions about my medical care (not related to the COVID-19 vaccine).
□ Results of the T-Detect test supported my decisions about receiving a COVID-19 vaccine.
□ I did nothing with the results.
□ Other

If Q8 = Other
Q9. Please describe what you did with the results.
___________________________

Q10. On a scale of 1-5, how useful were the results of the T-Detect COVID test?
□ 5: Extremely useful
□ 4: Very useful
□ 3: Somewhat useful
□ 2: Slightly useful
□ 1: Not at all useful

If Q4 = My healthcare provider recommended the test
Q11. To the best of your knowledge, why did your healthcare provider recommend the T-Detect COVID Test for you? (Select all that apply)
□ My healthcare provider wanted to understand my prior COVID infection status to support decisions about my daily activities.
□ My healthcare provider wanted to understand my prior COVID infection status to support a medical care decision (not related to the vaccine).
□ To aid in my decision regarding receiving a COVID vaccine
□ My healthcare provider wanted to determine whether my "Long-COVID" symptoms are related to a past infection that had not previously been positively diagnosed.
□ My healthcare provider wanted to determine whether I had a detectable immune (T-cell) response following my prior COVID exposure.
□ My healthcare provider wanted to determine whether I had a detectable immune (T-cell) response following my prior COVID vaccine.
□ Other

If Q11 = Other
Q12. Please describe the other reason your healthcare provider recommended the T-Detect COVID Test?
___________________________

Q13. Did you discuss the results of the T-Detect COVID test with your healthcare provider?
□ Yes
□ I have not discussed the results with my healthcare provider yet, but I do plan to.
□ I have not discussed the results with my healthcare provider and do not plan to.

If Q13 = Yes
Q14. What did your healthcare provider do with the results of the T-Detect COVID test? (Select all that apply)
□ My healthcare provider assisted me with decisions about my daily activities.
□ My healthcare provider assisted me with decisions about my medical care (not related to the COVID-19 vaccine).
□ My healthcare provider assisted me with decisions about receiving a COVID-19 vaccine.
□ My healthcare provider did not make any decisions based on the test results.
□ Other

If Q14 = Other
Q15. Please describe what your healthcare provider did with the results.
___________________________

Q16. On a scale of 1-5, how useful were the results of the T-Detect COVID test in the context of why you or your healthcare provider ordered the test?
□ 5: Extremely useful
□ 4: Very useful
□ 3: Somewhat useful
□ 2: Slightly useful
□ 1: Not at all useful

IRB Approved at the
Protocol Level
Sep 27, 2021

COVID-19 Diagnosis, Symptoms and Treatment

Q1. Have you been diagnosed with COVID-19?
□ Yes
□ No
If Q1 = No ⟶ Medical History Section

If Q1 = Yes
Q2. Were you diagnosed with COVID-19 after being vaccinated?
□ Yes
□ No
□ I have not been vaccinated
□ Prefer not to answer

Q3. How were you diagnosed with COVID-19? (Select all that apply)
□ I had a positive COVID-19 test (the most common are PCR or antigen tests)
□ I tested positive for COVID-19 antibodies
□ Other

If Q3 = I had a positive COVID-19 test (the most common are PCR or antigen tests)
Q4. What positive COVID-19 test did you have? (Select all that apply)
□ PCR
□ Antigen test
□ Other

If Q4 = PCR
Q5. What date did you have your PCR test done? (Month/Day/Year)

_______________
(MM/DD/YYYY)

Q6. What was the reason for testing? (Select all that apply)
□ I had symptoms of COVID-19
□ I was exposed to someone with symptoms of COVID-19
□ I was exposed to someone with a confirmed diagnosis of COVID-19
□ A healthcare provider told me to get tested for COVID-19
□ I was tested for an event or travel
□ It was part of routine screening for work or to have a medical procedure
□ Other ,

If Q6 = Other
Q7. Please describe the other reason for testing:
___________________________

IRB Approved at the
Protocol Level
Sep 27, 2021

If Q4 = Antigen test
Q8. What date did you have your antigen test done? (Month/Day/Year)

______________
(MM/DD/YYYY)

Q9. What was the reason for testing? (Select all that apply)
□ I had symptoms of COVID-19
□ I was exposed to someone with symptoms of COVID-19
□ I was exposed to someone with a confirmed diagnosis of COVID-19
□ A healthcare provider told me to get tested for COVID-19
□ I was tested for an event or travel
□ It was part of routine screening for work or to have a medical procedure
□ Other

If Q9 = Other
Q10. Please describe the other reason for testing:

_________________________

If Q4 = Other
Q11. Please describe the other COVID-19 test:

_________________________

Q12. What date did you have your other test done? (Month/Day/Year)

______________
(MM/DD/YYYY)

Q13. What was the reason for testing? (Select all that apply)
□ I had symptoms of COVID-19
□ I was exposed to someone with symptoms of COVID-19
□ I was exposed to someone with a confirmed diagnosis of COVID-19
□ A healthcare provider told me to get tested for COVID-19
□ I was tested for an event or travel
□ It was part of routine screening for work or to have a medical procedure
□ Other

If Q13 = Other
Q14. Please describe the other reason for testing:

_________________________

If Q3 = I tested positive for COVID-19 antibodies
Q15. What date did you have your antibody test done? ((MM/DD/YYYY)

______________
(MM/DD/YYYY)

If Q3 = Other
Q16. Please describe the other way you were diagnosed with COVID-19?

**Please answer the following questions regarding your first (or only) COVID-19 infection. We will ask about re-infections later in the questionnaire.**

Q17. Did you experience symptoms related to COVID-19?
□ Yes
□ No

If Q17 = Yes
Q18. Did you experience any of the following common symptoms related to COVID-19?
□ Fever
□ Chills
□ Cough
□ Shortness of breath or difficulty breathing
□ Chest pain or pressure
□ Fatigue
□ Muscle or body aches
□ Headache
□ Sore Throat
□ Runny or stuffy nose
□ None

Q19. Did you experience any of the following gastrointestinal symptoms related to COVID-19?
□ Nausea or vomiting
□ Abdominal pain
□ Diarrhea
□ Loss of appetite
□ None

Q20. Did you experience any of the following neurological symptoms related to COVID-19?
□ Loss of taste or smell
□ Trembling or shaking
□ Tingling or numbness
□ Dizziness
□ Sudden confusion
□ Slurred speech
□ Difficulty waking up
□ Difficulty sleeping
□ Difficulty concentrating or focusing
□ Inability to exercise or be active
□ Anxiety
□ Memory problems
□ None

Q21. Did you experience symptoms related to COVID-19 other than the ones described above?
___________________________

Q22. Approximately, when did your symptoms start (DD/MM/YYYY)?
_______________
(MM/DD/YYYY)

Q23. Approximately, when did your symptoms stop (DD/MM/YYYY)?
_______________
(MM/DD/YYYY)

Q24. How did your symptoms, when they were the most severe, affect your daily activities? Please select one of the following:
□ I was fully active and able to carry out normal routine without restriction
□ I was unable to do strenuous or intense physical activity but able to carry out light housework or office work
□ I was walking and capable of all self-care but unable to carry out any work activities; up and about more than 50% of waking hours
□ I was capable of only limited self-care; confined to bed or chair more than 50% of waking hours
□ I was unable to carry on any self-care and was confined to the bed or chair

Q25. How did your symptoms affect your productivity or effectiveness at work? Please select one of the following:
□ There were no changes to my normal productivity or effectiveness at work
□ I was productive or effective at work most of the time
□ I was productive or effective at work some of the time
□ I was unable to be productive or effective at work

Q26. How much work did you miss due to your COVID-19 symptoms?
□ None (0-1 Day)
□ Several Days (2-6 Days)
□ More than half the days (7-11 Days)
□ Almost Daily (12-14 Days)
□ I am unable to work/no longer employed as a result of my condition
□ Not Applicable

Q27. Did you go to the emergency department for your symptoms?
□ Yes
□ No

Q28. Were you hospitalized due to your symptoms?
□ Yes
□ No

If Q28 = Yes

IRB Approved at the
Protocol Level
Sep 27, 2021

Q29. When were you hospitalized (Month/Day/Year)?

______________
(MM/DD/YYYY)

Q30. How long were you hospitalized for?

_________________________

Q31. Were you cared for in the intensive care unit (ICU)?
□ Yes
□ No

Q32. Did you require any of the following? (Select all that apply)
□ Supplemental oxygen (by nasal cannula, face mask, etc.)
□ Ventilator
□ ECMO
□ None of the Above

Q33. Did you receive any of the following treatments? (Select all that apply)
□ Remdesivir
□ Steroids
□ Convalescent plasma
□ Monoclonal antibody therapy
□ Hydroxychloroquine
□ Azithromycin
□ Other
□ None of the Above

If Q33 = Remdesivir
Q34. When did you take remdesivir? If known, please provide the start and end date of treatment.

_________________________

If Q33 = Steroids
Q35. When did you take steroids? If known, please provide the start and end date of treatment.

_________________________

If Q33 = Convalescent plasma
Q36. How many times did you receive convalescent plasma?
□ 1
□ 2
□ 3
□ 4
□ More than 4 times

Q36 will determine how many q's below the participant receives.
Q37. When did you receive convalescent plasma (Month/Day/Year)?

______________
(MM/DD/YYYY)

Q38. When did you receive convalescent plasma (Month/Day/Year)?

______________
(MM/DD/YYYY)

Q39. When did you receive convalescent plasma (Month/Day/Year)?

______________
(MM/DD/YYYY)

Q40. When did you receive convalescent plasma (Month/Day/Year)?

______________
(MM/DD/YYYY)

If Q36 = More than 4 times
Q41. Please describe the number of times and when you received convalescent plasma (Month/Day/Year)?

_________________________

If Q33 = Monoclonal antibody therapy
Q42. What monoclonal antibody did you receive (Month/Day/Year)?
□ Bamlanivimab (LY-CoV555)
□ Bamlanivimab + etesevimab
□ Casirivimab and imdevimab (REGN-COV2)
□ Infliximab (Remicade)
□ Other
□ Not sure

Q43. Please describe any other monoclonal antibody treatment you received:
_________________________

Q44. When did you receive monoclonal antibodies?

______________
(MM/DD/YYYY)

If Q33 = Hydroxychloroquine
Q45. When did you start taking hydroxychloroquine (Month/Day/Year)?

______________
(MM/DD/YYYY)

Q46. When did you stop taking hydroxychloroquine (Month/Day/Year)?

______________
(MM/DD/YYYY)

If Q33 = Azithromycin

Q47. When did you start taking azithromycin (Month/Day/Year)?

______________
(MM/DD/YYYY)

Q48. When did you stop taking azithromycin (Month/Day/Year)?

______________
(MM/DD/YYYY)

If Q33 = Other
Q49. Please describe any other treatments you received for COVID-19 and the approximate dates you received them.

_________________________

Q50. Have you recovered from COVID-19?
□ Yes
□ No
<u>If Q50 = No → Long Term Complications from COVID-19 Section</u>

If Q50 = Yes
Q51. How do you know you recovered from COVID-19? (Select all that apply)
□ My healthcare provider cleared me
□ I had a negative PCR or antigen test
□ I no longer have symptoms
□ Other

If Q51 = Other
Q52. Please describe how you know you recovered from COVID-19?

_________________________

Q53. When were you told you recovered from COVID-19 (Month/Day/Year)?

______________
(MM/DD/YYYY)

If Q50 = Yes
Q51. Have you been reinfected with COVID-19 after recovering from COVID-19?
□ Yes
□ No

If Q51 = Yes
Repeat questions Q2. – Q51
**<u>Please answer the following questions regarding your second COVID-19 infection.</u>**

<u>If Q51 = No → Long Term Complications from COVID-19 Section</u>

IRB Approved at the
Protocol Level
Sep 27, 2021

Long Term Complications from COVID-19

Q1. Since having COVID-19, have you been diagnosed with any of the following?
□ Heart attack
□ Stroke or mini stroke/TIA
□ Kidney problems
□ Deep vein thrombosis (DVT, "blood clot in leg")
□ Pulmonary embolism (PE, "blood clot in lung")
□ Other
□ None

If Q1 = Heart attack
Q2. When were you diagnosed with a heart attack? (Month/Day/Year)?

_______________
(MM/DD/YYYY)

Q3. Did your doctor think your diagnosis of a heart attack was related to COVID-19?
□ Yes
□ No
□ Not sure

If Q1 = Stroke or mini stroke/TIA
Q4. When were you diagnosed with a stroke or mini stroke/TIA? (Month/Day/Year)?

_______________
(MM/DD/YYYY)

Q5. Did your doctor think your diagnosis of a stroke or mini stroke/TIA was related to COVID-19?
□ Yes
□ No
□ Not sure

If Q1 = Kidney problems
Q6. When were you diagnosed with kidney problems? (Month/Day/Year)?

_______________
(MM/DD/YYYY)

Q7. Did your doctor think your diagnosis of kidney problems was related to COVID-19?
□ Yes
□ No
□ Not sure

If Q1 = Deep vein thrombosis (DVT, "clot in leg")
Q8. When were you diagnosed with deep vein thrombosis (DVT, "blood clot in leg")? (Month/Day/Year)?

_______________

IRB Approved at the
Protocol Level
Sep 27, 2021

(MM/DD/YYYY)

Q9. Did your doctor think your diagnosis of deep vein thrombosis (DVT, "blood clot in leg") was related to COVID-19?
□ Yes
□ No
□ Not sure

If Q1 = Pulmonary embolism (PE, "clot in lung")
Q10. When were you diagnosed with pulmonary embolism (PE, "blood clot in lung") (Month/Day/Year)?

_______________
(MM/DD/YYYY)

Q11. Did your doctor think your diagnosis of pulmonary embolism (PE, "blood clot in lung") was related to COVID-19?
□ Yes
□ No
□ Not sure

If Q1 = Other
Q12. Please describe your other condition(s):
___________________________

Q13. When were you diagnosed with other (Month/Day/Year)?

_______________
(MM/DD/YYYY)

Q14. Did your doctor think your diagnosis of other was related to COVID-19?
□ Yes
□ No
□ Not sure

Q15. Have you been diagnosed with a post-COVID condition by a healthcare provider (it may also have been called long COVID, long-haul COVID, post-acute COVID-19, long-term effects of COVID, or chronic COVID)?
□ Yes
□ No

If Q15 = No
Q16. Did you experience any new or ongoing health problems four or more weeks after your COVID-19 diagnosis?
□ Yes
□ No
If Q16 = No → <u>Medical History Section</u>

If Q15 or Q16 = Yes
Q17. Have you experienced any of the following respiratory and sinus symptoms since you had COVID-19?
□ Difficulty breathing or shortness of breath
□ Persistent cough
□ Chest pain, pressure or tightness
□ Sneezing
□ Runny nose
□ Sore throat
□ Other
□ None of the above

If Q17 = Other
Q18. Please describe the other respiratory and sinus symptoms you have experienced since you had COVID-19.
___________________________

Q19. When did you begin experiencing these symptoms? (Please mark when you first experienced the respiratory and sinus symptoms since being diagnosed with COVID-19, in the first 4 weeks, then months, as applicable.)
□ Week 1
□ Week 2
□ Week 3
□ Week 4
□ Month 2
□ Month 3
□ Month 4
□ Month 5
□ Month 6
□ Month 7
□ Month 8
□ Month 9
□ Month 10
□ Month 11
□ Month 12

Q20. Have you experienced extreme tiredness or fatigue since you had COVID-19?
□ Yes
□ No

Q21. When did you begin experiencing these symptoms? (Please mark when you first experienced extreme tiredness or fatigue since being diagnosed with COVID-19, in the first 4 weeks, then months, as applicable.)
□ Week 1
□ Week 2
□ Week 3

□ Week 4
□ Month 2
□ Month 3
□ Month 4
□ Month 5
□ Month 6
□ Month 7
□ Month 8
□ Month 9
□ Month 10
□ Month 11
□ Month 12

Q22. Have you experienced issues with brain fog (inability to focus, think clearly, plan, process, understand, and maintain a coherent stream of thought; abnormally slow or fast thoughts) since you had COVID-19?
□ Yes
□ No

Q23. When did you begin experiencing these symptoms? (Please mark when you first experienced brain fog symptoms since being diagnosed with COVID-19, for the first 4 weeks, then months, as applicable.)
□ Week 1
□ Week 2
□ Week 3
□ Week 4
□ Month 2
□ Month 3
□ Month 4
□ Month 5
□ Month 6
□ Month 7
□ Month 8
□ Month 9
□ Month 10
□ Month 11
□ Month 12

Q24. Have you experienced any of the following gastrointestinal issues since you had COVID-19?
□ Diarrhea
□ Abdominal pain
□ Constipation
□ Vomiting
□ Loss of appetite
□ None of the above

Q25. When did you begin experiencing these symptoms? (Please mark when you first experienced the gastrointestinal issues since being diagnosed with COVID-19, in the first 4 weeks, then months, as applicable.)
□ Week 1
□ Week 2
□ Week 3
□ Week 4
□ Month 2
□ Month 3
□ Month 4
□ Month 5
□ Month 6
□ Month 7
□ Month 8
□ Month 9
□ Month 10
□ Month 11
□ Month 12

Q26. Have you experienced any of the following headache or related issues since you had COVID-19?
□ Headaches, at the base of the skull
□ Headaches, in the temples
□ Headaches, behind the eyes
□ Headaches, diffuse (entire brain)
□ Headaches/pains after mental exertion
□ Sensation of brain warmth
□ Sensation of brain pressure
□ Migraines
□ Stiff neck
□ Other
□ None of the above

If Q26 = Other
Q27. Please describe the other headache or related issues you have experienced since you had COVID-19.
___________________________

Q28. When did you begin experiencing these symptoms? (Please mark when you first experienced headache or related issues since being diagnosed with COVID-19, in the first 4 weeks, then months, as applicable.)
□ Week 1
□ Week 2
□ Week 3
□ Week 4

□ Month 2
□ Month 3
□ Month 4
□ Month 5
□ Month 6
□ Month 7
□ Month 8
□ Month 9
□ Month 10
□ Month 11
□ Month 12

Q29. Have you experienced any of the following cardiovascular issues since you had COVID-19?
□ Tachycardia (high heart rate)
□ Bradycardia (low heart rate)
□ Heart palpitations (Feeling like your heart is racing, thumping or skipping beats)
□ Abnormally high blood pressure
□ Fainting
□ Blood clots
□ None of the above

Q30. When did you begin experiencing these symptoms? (Please mark when you first experienced the cardiovascular issues since being diagnosed with COVID-19, in the first 4 weeks, then months, as applicable.)
□ Week 1
□ Week 2
□ Week 3
□ Week 4
□ Month 2
□ Month 3
□ Month 4
□ Month 5
□ Month 6
□ Month 7
□ Month 8
□ Month 9
□ Month 10
□ Month 11
□ Month 12

Q31. Have you experienced any of the following muscle and joint issues since you had COVID-19?
□ Muscle spasms
□ Muscle aches
□ Joint pain

□ Bone aches
□ None of the above

Q32. When did you begin experiencing these symptoms? (Please mark when you first experienced the muscle and joint issues since being diagnosed with COVID-19, in the first 4 weeks, then months, as applicable.)
□ Week 1
□ Week 2
□ Week 3
□ Week 4
□ Month 2
□ Month 3
□ Month 4
□ Month 5
□ Month 6
□ Month 7
□ Month 8
□ Month 9
□ Month 10
□ Month 11
□ Month 12

Q33. Which of the following neurological sensation symptoms have you first experienced since you had COVID-19?
□ Skin sensations: burning, tingling, or itchiness without rash
□ Numbness/loss of sensation
□ Coldness
□ Tingling/prickling pins and needles
□ Electrical zaps/electrical shock sensation
□ Facial paralysis
□ Sensation of facial pressure/numbness
□ Weakness
□ None of the above

Q34. When did you begin experiencing these symptoms? (Please mark when you first experienced the neurological sensation symptoms since being diagnosed with COVID-19, in the first 4 weeks, then months, as applicable.)
□ Week 1
□ Week 2
□ Week 3
□ Week 4
□ Month 2
□ Month 3
□ Month 4
□ Month 5
□ Month 6

□ Month 7
□ Month 8
□ Month 9
□ Month 10
□ Month 11
□ Month 12

Q35. Have you experienced any of the following memory related symptoms since you had COVID-19?
□ Short-term memory loss
□ Long-term memory loss
□ Not being able to make new memories
□ Forgetting how to do routine tasks
□ Other
□ None of the above

If Q35 = Other
Q36. Please describe the other memory symptoms you have experienced since you had COVID-19.
___________________________

Q37. When did you begin experiencing these symptoms? (Please mark when you first experienced memory symptoms since being diagnosed with COVID-19, in the first 4 weeks, then months, as applicable.)
□ Week 1
□ Week 2
□ Week 3
□ Week 4
□ Month 2
□ Month 3
□ Month 4
□ Month 5
□ Month 6
□ Month 7
□ Month 8
□ Month 9
□ Month 10
□ Month 11
□ Month 12

Q38. Have you experienced any sleep issues since you had COVID-19?
□ Yes
□ No

If Q38 = Yes
Q39. Which of the following sleep issues have you experienced since you had COVID-19?

IRB Approved at the
Protocol Level
Sep 27, 2021

□ Lucid dreams, vivid dreams, or nightmares
□ Insomnia
□ Night sweats
□ Restless leg syndrome
□ Sleep apnea
□ Other

If Q39 = Other
Q40. Please describe the other sleep issues you have experienced since you had COVID-19.

___________________________

Q41. When did you begin experiencing these symptoms? (Please mark when you first experienced sleep issues since being diagnosed with COVID-19, in the first 4 weeks, then months, as applicable.)
□ Week 1
□ Week 2
□ Week 3
□ Week 4
□ Month 2
□ Month 3
□ Month 4
□ Month 5
□ Month 6
□ Month 7
□ Month 8
□ Month 9
□ Month 10
□ Month 11
□ Month 12

Q42. Have you experienced any of the following temperature issues since you had COVID-19?
□ Elevated temperature (98.8-100.4 degrees Fahrenheit)
□ Fever (100.4 degrees Fahrenheit)
□ Low temperature
□ Chills/flushing/sweats
□ Heat intolerance
□ Temperature lability (quick swings in and out of fever or elevated temperature)
□ None of the above

Q43. When did you begin experiencing these symptoms? (Please mark when you first experienced the temperature issues since being diagnosed with COVID-19, in the first 4 weeks, then months, as applicable.)
□ Week 1
□ Week 2
□ Week 3
□ Week 4

IRB Approved at the
Protocol Level
Sep 27, 2021

□ Month 2
□ Month 3
□ Month 4
□ Month 5
□ Month 6
□ Month 7
□ Month 8
□ Month 9
□ Month 10
□ Month 11
□ Month 12

Q44. Have you experienced any of the following skin and allergy symptoms since you had COVID-19?
□ Itchy skin
□ Peeling skin
□ COVID toes (discoloration, swelling, painful, or blistering toes)
□ New allergies (food, chemical, environmental, etc.)
□ Skin rashes
□ Other
□ None of the above

If Q44 = Other
Q45. Please describe the other skin and allergy symptoms you have experienced since you had COVID-19.

____________________________

Q46. When did you begin experiencing these symptoms? (Please mark when you first experienced the skin and allergy symptoms since being diagnosed with COVID-19, in the first 4 weeks, then months, as applicable.)
□ Week 1
□ Week 2
□ Week 3
□ Week 4
□ Month 2
□ Month 3
□ Month 4
□ Month 5
□ Month 6
□ Month 7
□ Month 8
□ Month 9
□ Month 10
□ Month 11
□ Month 12

Q47. Have you experienced any changes to your sense of smell or taste since you had COVID-19?
□ Yes
□ No

If Q47 = Yes
Q48. Which of the following sense of smell or taste issues have you experienced since you had COVID-19?
□ Loss of smell
□ Phantom or unusual smells
□ Heightened sense of smell
□ Loss of taste
□ Phantom or unusual tastes
□ Heightened sense of taste
□ Altered sense of taste

Q49. When did you begin experiencing these symptoms? (Please mark when you first experienced sense of smell or taste symptoms since being diagnosed with COVID-19, in the first 4 weeks, then months, as applicable.)
□ Week 1
□ Week 2
□ Week 3
□ Week 4
□ Month 2
□ Month 3
□ Month 4
□ Month 5
□ Month 6
□ Month 7
□ Month 8
□ Month 9
□ Month 10
□ Month 11
□ Month 12

Q50. Have you experienced any changes in your menstrual period cycles since you had COVID-19?
□ Yes
□ No
□ Not Applicable

Q51. Have you experienced any changes in your mood since you had COVID-19?
□ Yes
□ No

Q52. How severe were/are your symptoms over the course of the following weeks/months? Please select the most severe within each time period.

Q53. Week 1
□ No symptoms
□ Very mild
□ Mild
□ Moderate
□ Severe
□ Very severe

Q54. Week 2
□ No symptoms
□ Very mild
□ Mild
□ Moderate
□ Severe
□ Very severe

Q55. Week 3
□ No symptoms
□ Very mild
□ Mild
□ Moderate
□ Severe
□ Very severe

Q56. Week 4
□ No symptoms
□ Very mild
□ Mild
□ Moderate
□ Severe
□ Very severe

Q57. Month 2
□ No symptoms
□ Very mild
□ Mild
□ Moderate
□ Severe
□ Very severe

Q58. Month 3
□ No symptoms
□ Very mild

IRB Approved at the
Protocol Level
Sep 27, 2021

□ Mild
□ Moderate
□ Severe
□ Very severe

Q59. Month 4
□ No symptoms
□ Very mild
□ Mild
□ Moderate
□ Severe
□ Very severe

Q60. Month 5
□ No symptoms
□ Very mild
□ Mild
□ Moderate
□ Severe
□ Very severe

Q61. Month 6
□ No symptoms
□ Very mild
□ Mild
□ Moderate
□ Severe
□ Very severe

Q62. Month 7
□ No symptoms
□ Very mild
□ Mild
□ Moderate
□ Severe
□ Very severe

Q63. Month 8
□ No symptoms
□ Very mild
□ Mild
□ Moderate
□ Severe
□ Very severe

Q64. Month 9

□ No symptoms
□ Very mild
□ Mild
□ Moderate
□ Severe
□ Very severe

Q65. Month 10
□ No symptoms
□ Very mild
□ Mild
□ Moderate
□ Severe
□ Very severe

Q66. Month 11
□ No symptoms
□ Very mild
□ Mild
□ Moderate
□ Severe
□ Very severe

Q67. Month 12
□ No symptoms
□ Very mild
□ Mild
□ Moderate
□ Severe
□ Very severe

Q68. Please describe your experience with any relapsing symptoms. (Select all that apply)
□ I have not experienced relapses
□ My relapses happen in a regular pattern
□ My relapses happen in an irregular pattern
□ My relapses happen in response to a trigger (stress, alcohol, exertion, etc.)
□ My relapses are becoming shorter/easier over time
□ My relapses are becoming longer/more difficult over time
□ My relapse severity has stayed the same
□ My symptoms have slowly gotten better over time
□ My symptoms have stayed the same over time
□ My symptoms have slowly worsened over time
□ I got worse rapidly
□ I got better rapidly

IRB Approved at the
Protocol Level
Sep 27, 2021

Post-COVID-19 Functional Status

Q1. Can you live alone without any assistance from another person? (e.g. independently being able to eat, walk, use the toilet and manage routine daily hygiene)
□ Yes
□ No (if no, skip to Q9)

Q2. Are there duties/activities at home or at work which you are no longer able to perform yourself?
□ Yes (if yes, skip to Q5)
□ No

Q3. Do you suffer from symptoms, pain, depression or anxiety?
□ Yes
□ No (if no, skip to Q5)

Q4. Do you need to avoid or reduce duties/activities or spread these over time?
□ Yes
□ No

Q5. Over the past two weeks, how often did you have problems with productivity or effectiveness at work?
□ Not at all (0-1 Day)
□ Several Days (2-6 Days)
□ More than half the days (7-11 Days)
□ Almost Daily (12-14 Days)
□ Not Applicable

If Q5 does not = Not Applicable
Q6. Are these issues new since testing positive for COVID-19, or were they present before COVID-19?
□ These issues are new
□ I had these issues before COVID-19

Q7. Over the past two weeks, how often did you have problems with absenteeism
□ Not at all (0-1 Day)
□ Several Days (2-6 Days)
□ More than half the days (7-11 Days)
□ Almost Daily (12-14 Days)
□ Not Applicable

If Q7 does not = Not Applicable
Q8. Are these issues new since testing positive for COVID-19, or were they present before COVID-19?
□ These issues are new
□ I had these issues before COVID-19

Q9. How much are you currently affected in your everyday life by COVID-19? Please indicate which one of the following statements applies to you most.

□ I have no limitations in my everyday life and no symptoms, pain, depression, or anxiety.

□ I have negligible limitations in my everyday life as I can perform all usual duties/activities, although I still have persistent symptoms, pain, depression, or anxiety.

□ I suffer from limitations in my everyday life as I occasionally need to avoid or reduce usual duties/activities or need to spread these over time due to symptoms, pain, depression or anxiety. I am, however, able to perform all activities without any assistance.

□ I suffer from limitations in my everyday life as I am not able to perform all usual duties/activities due to symptoms, pain, depression, or anxiety. I am, however, able to take care of myself without any assistance

□ I suffer from severe limitations in my everyday life: I am not able to take care of myself and therefore I am dependent on nursing care and/or assistance from another person due to symptoms, pain, depression or anxiety.

Past Medical History

Q1. Please list all of your current medications (prescribed, over the counter and supplements):
___________________________

Q2. Do you currently take any medication that suppress your immune system, such as steroids or other immunosuppressants)?
□ Yes
□ No

If Q2 Yes
Q3. Please list the medication(s):
___________________________

Q4. Have you been diagnosed with any of the following medical conditions? (Select all that apply)
□ Diabetes (Type 1)
□ Diabetes (Type 2)
□ Hypertension
□ Heart disease
□ Lung disease
□ Asthma
□ Stroke
□ Obesity
□ None

Q5. When were you diagnosed (MM/YYYY)?
_____________
(MM//YYYY)

Q6. Have you ever been diagnosed with any of the following autoimmune diseases?
□ Rheumatoid Arthritis
□ Type 1 Diabetes (i.e. started when you were younger and requiring insulin)
□ Celiac disease
□ Lupus
□ Ulcerative Colitis
□ Crohn's disease
□ Multiple Sclerosis
□ Scleroderma
□ Sjögren's disease
□ Primary biliary cirrhosis
□ Primary sclerosing cholangitis
□ Psoriasis
□ Hashimoto's thyroiditis
□ Autoimmune hepatitis
□ Addison's disease

IRB Approved at the
Protocol Level
Sep 27, 2021

□ Other
□ None

If Q6=Other
Q7. Please list your other diagnosed autoimmune diseases:

___________________________

Q8. When were you diagnosed (MM/YYYY)?

_____________
(MM//YYYY)

If Q6. Does not = None, i.e. any other response is selected
Q9. Do you currently take any medication to manage your autoimmune disease (such as a steroid or immunosuppressant)?
□ Yes
□ No

If Q9 =Yes
Q10. Please list your current medications to manage your autoimmune disease:

___________________________

Q11. Have you ever been diagnosed with cancer?
□ Yes
□ No

If Q11 =Yes
Q12. Please list your diagnosed cancer(s):

___________________________

Q13. When were you diagnosed (MM/YYYY)?

_____________
(MM//YYYY)

Q14. Are you currently taking any medication or receiving treatment for your cancer?
□ Yes
□ No

If Q14 =Yes
Q15. Please list your current medications or treatment for your cancer:

___________________________

Q16. Have you ever received a bone marrow, cord blood or other stem cell transplant as a treatment for your cancer?
□ Yes
□ No

IRB Approved at the
Protocol Level
Sep 27, 2021

Q17. Have you ever been diagnosed with any of the following infectious diseases?
□ Chickenpox
□ Diphtheria
□ E. coli
□ Giardiasis
□ HIV/AIDS
□ Hepatitis B
□ Hepatitis C
□ Infectious mononucleosis
□ Lyme disease
□ Malaria
□ Measles
□ Meningitis
□ Mumps
□ Pelvic Inflammatory Disease (PID)
□ Poliomyelitis (polio)
□ Pneumonia
□ Rocky mountain spotted fever
□ Rubella (German measles)
□ Severe acute respiratory syndrome (SARS)
□ Sexually transmitted diseases
□ Shingles (herpes zoster)
□ Tetanus
□ Toxic shock syndrome
□ Tuberculosis
□ Viral hepatitis
□ West Nile virus
□ Whooping cough (pertussis)
□ Other
□ None

If Q17 = Sexually transmitted diseases
Q18. What sexually transmitted disease have you been diagnosed with?
□ Bacterial Vaginosis
□ Chlamydia
□ Gonorrhea
□ Herpes
□ Human papillomavirus (HPV)
□ Syphilis
□ Trichomoniasis
□ Other
□ Prefer not to answer

If Q18=Other

Q19. Please list your other diagnosed sexually transmitted diseases:

___________________________

If Q7=Other

Q20. Please list your other diagnosed infectious diseases:

___________________________

Q21. When were you diagnosed (MM/YYYY)?

_____________

(MM//YYYY)

If Q17. Does not = None, i.e. any other response is selected

Q22. Do you currently take any medication to manage your infectious disease (such as a steroid or immunosuppressant)?

□ Yes
□ No

If Q22 = Yes

Q23. Please list your current medications to manage your infectious disease:

___________________________

Family Medical History

Please complete the following family medical history for your first-degree and second-degree relatives as defined below:

- A first-degree relative is defined as a close blood relative which includes your parents, full siblings, or children.
- A second-degree relative is defined as a blood relative which includes your grandparents, grandchildren, aunts, uncles, nephews, nieces or half-siblings.

Q1. Do you have a first-degree relative who has been diagnosed with any of the following medical conditions? Please select all that apply:
□ Diabetes (Type 1)
□ Diabetes (Type 2)
□ Hypertension
□ Heart Disease
□ Lung Disease
□ Asthma
□ Stroke
□ Obesity
□ Other
□ None
□ Not sure

If Q1 = Other
Q2. Please list the other diagnosed medical conditions:
___________________________

Q3. Do you have a second-degree relative who has been diagnosed with any of the following medical conditions? Please select all that apply:
□ Diabetes (Type 1)
□ Diabetes (Type 2)
□ Hypertension
□ Heart Disease
□ Lung Disease
□ Asthma
□ Stroke
□ Obesity
□ Other
□ None
□ Not sure

If Q3 = Other
Q4. Please list the other diagnosed medical conditions:
___________________________

Q5. Do you have a first-degree relative who has been diagnosed with any of the following autoimmune diseases?
□ Rheumatoid Arthritis
□ Type 1 Diabetes (i.e. started when you were younger and requiring insulin)
□ Celiac disease
□ Lupus
□ Ulcerative Colitis
□ Crohn's disease
□ Multiple Sclerosis
□ Scleroderma
□ Sjögren's disease
□ Primary biliary cirrhosis
□ Primary sclerosing cholangitis
□ Psoriasis
□ Hashimoto's thyroiditis
□ Autoimmune hepatitis
□ Addison's disease
□ Other
□ None
□ Not sure

If Q5=Other
Q6. Please list the other diagnosed autoimmune diseases:
___________________________

Q7. Do you have a second-degree relative who has been diagnosed with any of the following autoimmune diseases?
□ Rheumatoid Arthritis
□ Type 1 Diabetes (i.e. started when you were younger and requiring insulin)
□ Celiac disease
□ Lupus
□ Ulcerative Colitis
□ Crohn's disease
□ Multiple Sclerosis
□ Scleroderma
□ Sjögren's disease
□ Primary biliary cirrhosis
□ Primary sclerosing cholangitis
□ Psoriasis
□ Hashimoto's thyroiditis
□ Autoimmune hepatitis
□ Addison's disease
□ Other
□ None
□ Not sure

If Q7=Other
Q8. Please list the other diagnosed autoimmune diseases:

___________________________

Q9. Do you have a first-degree relative who has diagnosed with cancer?
□ Yes
□ No

If Q9 = Yes
Q10. Please list the diagnosed cancer(s):

___________________________

Q11. Do you have a second-degree relative who has diagnosed with cancer?
□ Yes
□ No

If Q11 = Yes
Q12. Please list the diagnosed cancer(s):

___________________________
